# Pleiotropic effects of statins on ischemic heart disease: a Mendelian Randomization study in the UK Biobank

**DOI:** 10.1101/2020.01.14.20017400

**Authors:** CM Schooling, JV Zhao, SL Au Yeung, GM Leung

**Affiliations:** School of Public Health, Li Ka Shing Faculty of Medicine, The University of Hong Kong, Hong Kong, China; City University of New York, Graduate School of Public Health and Health Policy, New York, NY, USA

**Author notes:** Corresponding Author: CM Schooling, PhD. 1/F, Patrick Manson Building (North Wing), 7 Sassoon Road, Pokfulam, Hong Kong.

## Abstract

**Objectives:** Statins appear to have pleiotropic effects. We examined whether specifically statins, of the major lipid modifiers, operate on ischemic heart disease (IHD) via testosterone. As a validation, we assessed whether a drug that unexpectedly likely increases IHD also operates via testosterone.

**Design:** A sex-specific univariable and multivariable Mendelian randomization study

**Setting:** A large, population-based cohort study recruited in the UK from 2006-10, the UK Biobank

**Participants:** 179918 men with 25410 IHD cases and 212080 women with 12511 IHD cases

**Main Outcome measures:** Testosterone and IHD

**Results:** Of the three lipid modulations considered, statins, proprotein convertase subtilisin/kexin type 9 (PCSK9) inhibitors and ezetimibe, only genetically predicted statin use in men affected testosterone (−0.15 effect size testosterone per effect size lower (of low-density lipoprotein cholesterol), 95% confidence interval (CI) −0.23 to −0.06). The genetically predicted effect of statin use on IHD in specifically men was partially mediated by testosterone (odds ratio (OR) 0.55 per effect size lower (low-density lipoprotein cholesterol), 95% CI 0.38 to 0.79, compared to OR 0.73, 95% CI 0.46 to 1.11 after allowing for testosterone). The estimate for the effect of genetically predicted statin use, independent of testosterone, was very similar in women, giving overall meta-analyzed OR 0.72, 95% CI 0.57 to 0.90 per effect size lower of low-density lipoprotein cholesterol. The genetically predicted effect of anakinra use also affected testosterone (0.022 per effect size (of IL-1Ra), 95% CI 0.01 to 0.04), and increased IHD in men.

**Conclusions:** Statins may partially operate via testosterone in men, which may contribute to sex-specific pleiotropic effects. Anakinra operating by testosterone may also explain its unexpected effects. Our findings could facilitate the development of new interventions for cardiovascular diseases as well as highlighting the importance of sex-specific investigations and possibly treatments.

**Section 1: What is already known on this topic:** Statins appear to have pleiotropic effects on cardiovascular disease. Whether such effects exist and why they should occur is unclear, but could be highly relevant to the prevention and treatment of the leading cause of death.

**Section 2: What this study adds:** Our study shows that statins have similar protective effects on ischemic heart disease via low-density lipoprotein cholesterol in men and women, but unlike other major lipid modifiers statins have an additional effect specific to men via testosterone, while any harms of anakinra in men may operate by a similar mechanism. Our findings highlighting the possibility of sex-specific causes of cardiovascular disease and the need for sex-specific investigations, prevention and treatment.

## Background

Statins are the first-line lipid modifier for reducing cardiovascular morbidity and mortality [1, 2]. Statins have revolutionized the prevention and treatment of cardiovascular disease, and inspired the development of a range of effective interventions targeting the reduction of low-density lipoprotein (LDL)-cholesterol. Statins have long been suspected of having additional beneficial effects beyond lipid modulation [3], such as on inflammation [3], another potential target for reducing cardiovascular disease [4]. Meta-analysis of randomized controlled trials (RCTs) suggests statins are more effective at reducing mortality than proprotein convertase subtilisin/kexin type 9 (PCSK9) inhibitors or ezetimibe [5, 6]. However, these findings may be more apparent than real, stemming from differences in trial design, such as shorter duration of the PCSK9 inhibitor trials [6], the predominance of industry funded statin trials [7] or the difficulty of interpreting trials of “soft” events when the treatment affects diagnostically relevant criteria, i.e., lipid levels [8]. To investigate this anomaly, a previous study conducted a systematic agnostic scan of metabolic profile in a trial of statins compared to a PCSK9 inhibitor, which found few differences [9]. While characterization of the metabolic effects of statins suggested extensive effects on lipids and fatty acids [10]; these investigations were not able to include a factor which has previously been proposed as contributing to statin’s effectiveness, i.e., effects on male hormones [11], although questions have been raised as to whether statins are as effective in women as men [12]. However, meta-analysis of the available trial evidence suggests similar relative benefits of LDL-cholesterol reduction by statins for men and women [13], although the trials mainly concern men (73.2%) which may preclude detection of important sex differences. Men are also at substantially higher risk than women [14] giving larger absolute benefits in men than women at the same reduction in relative risk.

RCTs are not usually designed or powered to test mediating mechanisms. In addition, trials of statins on cardiovascular disease outcomes designed to be sex-specific are lacking. To assess a potential pathway by which statins might additionally operate, we used Mendelian randomization (MR), an observational study design that avoids confounding by taking advantage of the random allocation of genetic material at conception [15], here specifically the genetic variants corresponding to effects of use of lipid modifiers. This random allocation at conception also avoids selection bias as long as few deaths have occurred between randomization and recruitment due to exposure, outcome, or other causes, i.e., competing risk, of the outcome [16, 17]. So, here we focused on ischemic heart disease (IHD) [18], using the UK Biobank [19] to investigate whether testosterone mediated any of the effects of statins, PCSK9 inhibitors or ezetimibe on IHD in men or women using univariable and multivariable MR. As a further test, given some anti-inflammatories have also been shown to have the opposite effects on male hormones compared to statins, specifically the interleukin 1 receptor antagonist (IL-1Ra), anakinra [20], we assessed whether the genetic variants corresponding to effects of anakinra or tocilizumab use targeting interleukin 6 (IL-6 and IL-6R) had the mirror image pattern of effects on testosterone and IHD [4, 21, 22] to statins. Figure 1 illustrates the possible additional effects of statins, anakinra or tocilizumab on IHD via male hormones in the context of the well-established benefits of statins, PCSK9 inhibitors and ezetimibe acting via LDL-cholesterol and of anti-inflammatories in IHD.

**Figure 1:**
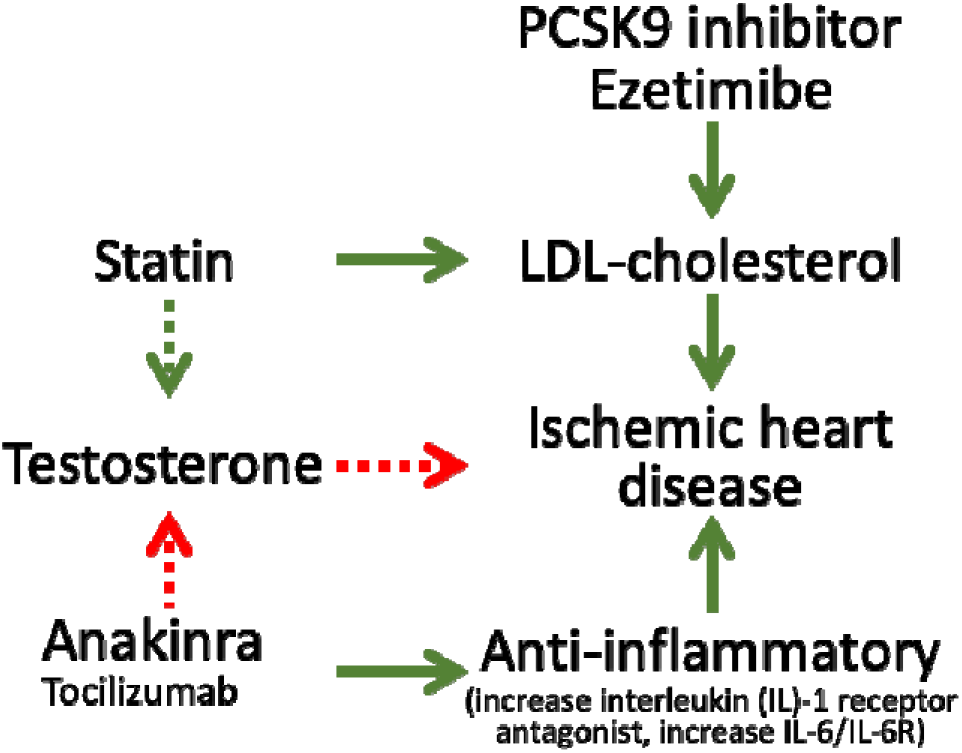
Directed acyclic graph showing the well-established protective effects of lipid modifiers and anti-inflammatories on IHD (solid lines) and possible additional pathways (dashed lines) investigated here. Green indicates a lowering effect, red indicates an increasing effect.

## Methods

### Genetic predictors of effects of lipid and interleukin modifier use

Established genetic variants corresponding to effects of statin, PCSK9 inhibitor and ezetimibe use were taken from published sources [23] which selected single nucleotide polymorphisms (SNPs) from genes encoding proteins of the targets of each lipid modifier (*HMGCR* for statins, *PCSK9* for PCSK9 inhibitors and *NCP1L1* for ezetimibe) that lowered LDL-cholesterol. Genetic effects of using statins, PCSK9 inhibitors and ezetimibe were expressed in sex-specific effect sizes of LDL-cholesterol reduction taken from the largest available GWAS summary statistics, i.e., the UK Biobank (http://www.nealelab.is/uk-biobank). The study was restricted to people of white British ancestry adjusted for the first 20 principal components, age, and age^2^. For each lipid modifier, we used independent (r^2^<0.05) SNPs most strongly associated with LDL-cholesterol, based on the 1000 Genomes catalog obtained from LDlink (https://ldlink.nci.nih.gov) in the main analysis, and then included all relevant SNPs in a sensitivity analysis, accounting for their correlations obtained from LDlink. Established genetic variants corresponding to anakinra and tocilizumab use and their effects on IL-1Ra and IL-6 respectively were also taken from published sources [21, 22].

### Sex-specific genetic predictors of testosterone

Strong (p-value<5×10^−8^), independent (r^2^<0.05), sex-specific genetic predictors of testosterone were extracted from a published genome wide association study (GWAS) based on the UK Biobank and replicated in three independent studies (CHARGE Consortium, Twins UK and EPIC-Norfolk) [24, 25]. Genetic associations with testosterone in this study were adjusted for genotyping chip/release of genetic data, age at baseline, fasting time and ten genetically derived principal components [24]. We used all 125 genetic variants given for bioavailable testosterone, hereafter testosterone, in men and all 254 genetic variants given for testosterone in women, as previously [26], because these had little correlation with sex hormone binding globulin (0.05 in men and 0.06 in women) [24].

### Sex-specific genetic associations with IHD

Sex-specific genetic associations with IHD were taken from the UK Biobank individual data after excluding those with inconsistent self-reported and genotyped sex, excess relatedness (more than 10 putative third-degree relatives), abnormal sex chromosomes (such as XXY), or poor-quality genotyping (heterozygosity or missing rate >1.5%). The sex-specific association with IHD were adjusted for the first 20 principal components, age, and assay array. IHD was based on self-report at baseline, subsequent health service encounter diagnoses (primary or secondary) of International Classification of Diseases (ICD) 9 410-4 or ICD10 I20-5 and death registration causes (primary or secondary) of ICD10 I20-5 up until December 2019.

### Statistical analysis

The F-statistic was used to assess instrument strength, obtained using an approximation (mean of square of SNP-exposure association divided by square of its standard error) [27]. A conventional threshold for the F-statistic is 10. SNPs with an F-statistic <10 were dropped.

Sex-specific estimates of the associations of genetically predicted exposures (i.e., genetically predicted effects of statin, PCSK9 inhibitor, ezetimibe, anakinra and tocilizumab use) with testosterone and IHD, as well as estimates of the associations of genetically predicted testosterone with IHD were obtained by combining SNP-specific Wald estimates (SNP on outcome divided by SNP on exposure) using inverse variance weighting (IVW) with multiplicative random effects [28]. Multivariable MR was used to assess sex-specific associations of genetically predicted exposures with IHD allowing for testosterone, accounting for correlations between SNPs on the same chromosome obtained from LDlink. In the multivariable MR, we estimated the Sanderson-Windmejier conditional F-statistic [29] to obtain a lower bound of the strength for each instrument conditional on the other exposure, and the Q statistics to asses pleiotropy, using the WSpiller/MVMR package [30].

### Sensitivity analysis

Where possible we used methods with different assumptions to assess the validity of the MR estimates from IVW, which assumes balanced pleiotropy. MR-Egger is valid as long as the instrument strength independent of direct effect assumption holds [31]. We also used a weighted median which gives valid estimates when more than 50% of information comes from valid SNPs [32]. However, for exposures instrumented by correlated SNPs we did not give the weighted median or MR-Egger estimates because of concerns about their interpretability [33].

All statistical analysis was conducted using R version 3.6.1 (The R Foundation for Statistical Computing, Vienna, Austria). The MendelianRandomization R package was used for the MR estimates. Estimates of genetic associations were taken from publicly available UK Biobank summary statistics, except the associations with IHD which were based on individual level genetic associations from the UK Biobank obtained under application #42468. All UK Biobank data was collected with fully informed consent.

### Patient and Public Involvement

Patients or the public were not involved in the design, or conduct, or reporting, or dissemination plans of our research

## Results

The six SNPs for the effects of statin use (rs12916, rs5909, rs10066707, rs17238484, rs2006760 and rs2303152) [23] were all correlated (r^2^>0.13). In the main analysis we used only the lead SNP, rs12916. Of the 7 SNPs for the effects of PCSK9 inhibitor use (rs11206510, rs2149041, rs7552841, rs10888897, rs2479394, rs2479409 and rs562556) [23], the three independently (r^2^<0.05) and most strongly associated with LDL-cholesterol (rs11206510, rs2149041 and rs7552841) were used in the main analysis. Of the 5 SNPs for the effects of ezetimibe use (rs10260606 (proxy of rs2073547), rs2300414, rs10234070, rs7791240, rs217386) [23], two SNPs (rs2300414 and rs10234070) were discarded because their F-statistic for LDL-cholesterol was <10. The remaining three SNPs were all correlated at r^2^>0.05. rs2073547 was used in the main analysis because it had the strongest association with LDL-cholesterol. Table S1 shows the associations with LDL-cholesterol by sex for the independent SNPs used to predict effects of statin, PCSK9 inhibitor and ezetimibe use. Table S2 shows the associations of the two SNPs for the effects of the use of the anti-inflammatory, anakinra, (rs6743376 and rs1542176 (r^2^<0.001)) with IL-1Ra and the associations of the SNP (rs7529229), used to predict effects of tocilizumab use, with IL-6.

There were 179,918 men with 25,410 cases of IHD and 212,080 women with 12,511 cases of IHD in the UK Biobank.

### Instrument strength

The F-statistics for SNPs used to genetically predict the effects of statin, PCSK9 and ezetimibe use were all >10 in men and women (Table S1), as were the F-statistics for the SNPs used to genetically predict the effects of anakinra and tocilizumab use (Table S2). The F-statistics for the 125 and 254 SNPs predicting testosterone in men and women were all greater than 10, with mean 128.6 and 83.3, respectively (Tables S3 and S4).

### Sex-specific associations of genetically predicted lipid modifiers with testosterone

Genetically predicted effects of statin use reduced testosterone in men but not women (Table 1). Genetically predicted effects of PCSK9 inhibitors or of ezetimibe did not affect testosterone in men or women (Table 1). Findings were similar in sensitivity analysis using a larger number of SNPs, where available (Table 1). PCSK9 inhibitors and ezetimibe were not investigated further, given the lack of association with testosterone in men and women.

**Table 1:**
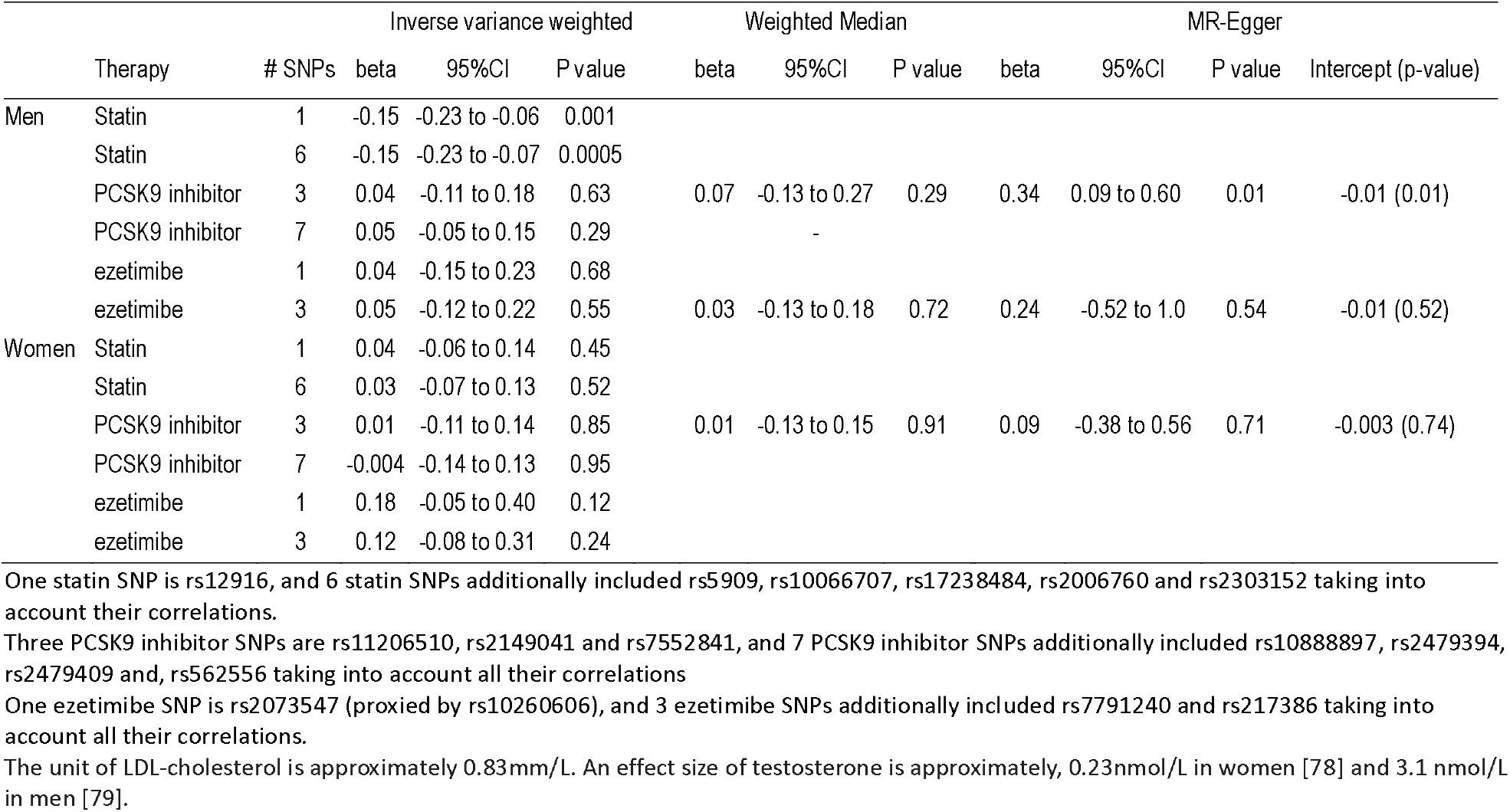
Sex-specific Mendelian randomization estimates for effects of genetically predicted statin, PCSK9 inhibitor and ezetimibe use (in effect sizes of LDL-cholesterol) on testosterone (effect size) in men and women using the UK Biobank summary statistics.

### Sex-specific associations of genetically predicted statin use and testosterone with IHD

Genetically predicted effects of statin use reduced both testosterone and the risk of IHD in men and possibly women (Table 2) using IVW. Genetically predicted testosterone was positively associated with IHD in men, but less clearly in women, with similar estimates using IVW, the weighted median and MR-Egger. MR-Egger intercepts did not suggest the IVW estimates were invalid, but had wider confidence intervals (Table 2)

**Table 2:** Mendelian randomization estimates for effects of genetically predicted statin use (effect sizes of LDL-cholesterol) and of testosterone (effect size) on IHD in men and women using the UK Biobank individual data.

|  | Exposure | # SNPs | Inverse variance weighted |  |  | Weighted Median |  |  | MR-Egger |  |  |  |
| --- | --- | --- | --- | --- | --- | --- | --- | --- | --- | --- | --- | --- |
|  |  |  | OR | 95%CI | P value | OR | 95%CI | P value | OR | 95%CI | P value | Intercept (p-value) |
| Men | Statin use | 1 | 0.55 | 0.38 to 0.79 | 0.001 |  |  |  |  |  |  |  |
|  | Statin use | 6 | 0.54 | 0.33 to 0.89 | 0.02 |  |  |  |  |  |  |  |
|  | Testosterone | 125 | 1.11 | 1.04 to 1.19 | 0.003 | 1.18 | 1.06 to 1.31 | 0.002 | 1.10 | 0.98 to 1.23 | 0.09 | 0.01 (0.84) |
| Women | Statin use | 1 | 0.87 | 0.59 to 1.27 | 0.46 |  |  |  |  |  |  |  |
|  | Statin use | 6 | 0.79 | 0.54 to 1.13 | 0.20 |  |  |  |  |  |  |  |
|  | Testosterone | 254 | 0.96 | 0.89 to 1.03 | 0.29 | 1.03 | 0.92 to 1.14 | 0.63 | 1.08 | 0.94 to 1.23 | 0.27 | -0.004 (0.05) |

One statin SNP is rs12916, and 6 statin SNPs additionally included rs5909, rs10066707, rs17238484, rs2006760 and rs2303152 taking into account all their correlations. The unit of LDL-cholesterol is approximately 0.83mm/L. An effect size of testosterone is approximately, 0.23nmol/L in women [78] and 3.1 nmol/L in men [79].

Considering genetically predicted effects of statin use together with genetically predicted testosterone, the conditional F-statistics were 58.2 (men) and 68.5 (women) for testosterone and 3.5 (men) and 6.8 (women) for effects of statin use. The Q statistics for instrument validity were significant (212.5 in men and 323.1 in women). Correspondingly, the multivariable MR-Egger intercept was significant in men and women, so we used the MR-Egger estimates. Multivariable associations of genetically predicted testosterone with IHD, adjusted for statin use, (Table 3) were similar to the univariable estimates for men and women (Table 2), but differed by sex (z-test p-value 0.01). For men the multivariable estimate for genetically predicted effects of statin use allowing for testosterone was attenuated (Table 3) compared to the univariable estimates (Table 2). As a result, the estimates for genetically predicted effects of statin use independent of testosterone were very similar for men and women (meta-analyzed odds ratio 0.72, 95% confidence interval 0.57 to 0.90).

**Table 3:** Multivariable Mendelian randomization estimates for effects of genetically predicted statin use (effect sizes of LDL-cholesterol) and of testosterone (effect size) together on IHD in men and women.

|  | Exposure | Instrumented by | Adjusted for | Inverse variance weighted |  |  | MR-Egger |  |  |  |
| --- | --- | --- | --- | --- | --- | --- | --- | --- | --- | --- |
|  |  |  |  | OR | 95%CI | P value | OR | 95% CI | p-value | Intercept p-value |
| Men | Statin use | 1 Statin SNP on LDL-cholesterol | Testosterone | 1.05 | 0.74 to 1.47 | 0.79 | 0.73 | 0.48 to 1.11 | 0.14 |  |
|  | Testosterone | 125 SNPs on testosterone | statin SNPs | 1.11 | 1.04 to 1.20 | 0.003 | 1.09 | 1.02 to 1.17 | 0.02 | 0.005 |
|  | Statin use | 6 Statin SNPs on LDL-cholesterol | Testosterone | 1.02 | 0.72 to 1.43 | 0.91 |  |  |  |  |
|  | Testosterone | 125 SNPs on testosterone | statin SNPs | 1.11 | 1.04 to 1.20 | 0.003 |  |  |  |  |
| Women | Statin use | 1 Statin SNP on LDL-cholesterol | Testosterone | 0.98 | 0.75 to 1.16 | 0.53 | 0.72 | 0.55 to 0.94 | 0.02 |  |
|  | Testosterone | 254 SNPs on testosterone | statin SNPs | 0.96 | 0.90 to 1.04 | 0.33 | 0.96 | 0.89 to 1.03 | 0.27 | 0.001 |
|  | Statin use | 6 Statin SNPs on LDL-cholesterol | Testosterone | 0.92 | 0.74 to 1.16 | 0.49 |  |  |  |  |
|  | Testosterone | 254 SNPs on testosterone | statin SNPs | 0.97 | 0.90 to 1.04 | 0.36 |  |  |  |  |

One statin SNP is rs12916, and 6 statin SNPs additionally included rs5909, rs10066707, rs17238484, rs2006760 and rs2303152 taking into account all their correlations. The unit of LDL-cholesterol is approximately 0.83mm/L. An effect size of testosterone is approximately, 0.23nmol/L in women [78] and 3.1 nmol/L in men [79]

### Sex-specific associations of genetically predicted anakinra and tocilizumab use with testosterone and IHD

Genetically predicted effects of anakinra use increased both the risk of IHD and testosterone in men but not women (Table 4). Genetically predicted effects of tocilizumab use were not clearly associated with testosterone in men or women (Table 4), so were not investigated further. Investigation of whether testosterone mediates the effect of anakinra use on IHD was not possible because sex-specific genetic associations of testosterone SNPs with IL-1Ra from suitably large GWAS are not available.

**Table 4:**
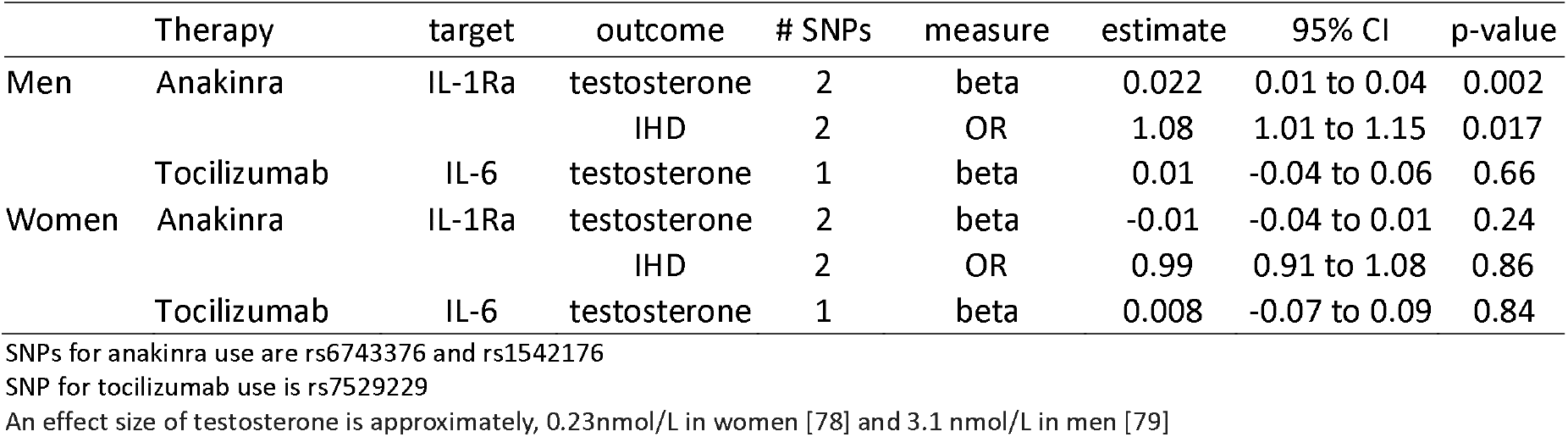
Mendelian randomization inverse variance weighted estimates for genetic effects of use of the anti-inflammatory anakinra raising IL-1Ra (effect size) [21] on testosterone (effect size) and ischemic heart disease and for genetic effects of use of tocilizumab raising IL-6 (log(pg/ml)) and IL-6R [22] on testosterone in men and women using the UK Biobank individual data.

## Discussion

Consistent with meta-analysis of RCTs this study provides genetic evidence that statins reduce testosterone in men [11], and adds by showing that statins partially operate on IHD in men only by reducing testosterone, as previously hypothesized [11, 34], while having similar protective effects in men and women independent of testosterone. Previous Mendelian randomization studies have shown lower testosterone associated with lower risk of IHD, particularly in men [35-37]. Conversely, consistent with an RCT [20], this study also provides genetic validation that the anti-inflammatory anakinra, targeting IL-1Ra, increases testosterone in men, and is consistent with a previous Mendelian randomization study showing anakinra increases IHD [22], but adds by showing why these associations might occur and that they may be specific to men.

A more marked association of testosterone with IHD in men than women (Table 2) is consistent with sex differences in biology, where testosterone is the main sex hormone in men and is much higher in men than in women. The associations of genetically predicted effects of statin, PCSK9 inhibitor or ezetimibe use on testosterone is consistent with the evidence available [11, 38, 39] and their mechanisms of action. Specifically, statins inhibit cholesterol synthesis, while PCSK9 inhibitors enable greater clearance of cholesterol, through increasing LDL-receptors, while ezetimibe reduces uptake of dietary cholesterol [1, 2]. However, some cells, such as Leydig cells, use *de novo* cholesterol synthesis to generate steroids, which can be reduced by statins [40]. Concerns about statins compromising androgen production pre-date the marketing of statins [41, 42]. Beneficial immunosuppressive effects of androgens in rheumatoid arthritis have long been known [43], making androgen reduction a plausible mode of action for therapies, such as anakinra, whose primary indication is rheumatoid arthritis.

These findings may seem counter-intuitive given the essential role of testosterone in masculinity and reproduction. However, in 2015 the Food and Drug Administration in the United States required labelling changes for all testosterone prescriptions to warn of the risk of heart attacks and stroke on testosterone, although no sufficiently large RCT of testosterone administration has been conducted to confirm these effects [44]. The Endocrine Society has also recommended caution in the use of testosterone [45]. Meta-analysis of RCTs suggests androgen deprivation therapy reduces all-cause mortality, but is too small to quantify effects on specific diseases beyond prostate cancer [46]. As such, our Mendelian randomization findings of the effects of testosterone have some consistency with the limited experimental evidence. In addition, our findings are consistent with well-established evolutionary biology theory, i.e. the Darwinian imperative of reproductive success even at the expense of longevity, possibly in a sex-specific manner, implies that central drivers of the reproductive axis, as well as androgen production and catabolism, and their environmental cues may be relevant to IHD [47, 48] (Figure 2) encompassing the relations tested here (Figure 1).

**Figure 2:**
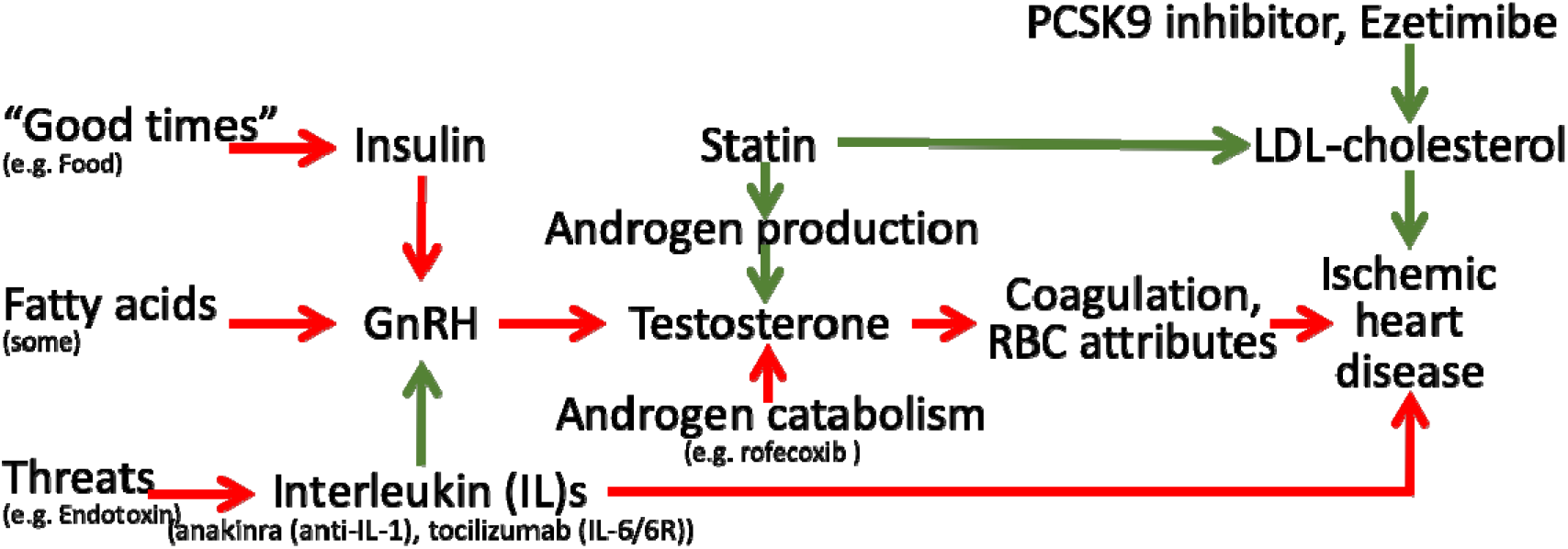
Schematic diagram showing the well-established protective effects of lipid modifiers on IHD (solid green lines) in the context of additional relevant pathways (green protective, red harmful) from an evolutionary biology perspective. Key: GnRH: gonadotropin releasing hormone, RBC: red blood cell, LDL: low density lipoprotein

Notably, upregulation of indicators of plentiful living conditions, such as insulin, appear to cause IHD, particularly in men [49], likely via gonadotropin releasing hormone (GnRH) [48]. Similarly, fatty acids may affect GnRH [50, 51]. In contrast, indicators of adversity, such as endotoxins promote an inflammatory response, involving interleukins, which suppresses the reproductive axis [52] and thereby testosterone [53], which may be reversed by anakinra possibly outweighing the benefits for IHD of suppressing inflammation [54]. Statins, in contrast reduce androgen production, while agents that suppress androgen catabolism, such as rofecoxib, have also had unexpectedly adverse effects on IHD [47]. Mechanisms by which androgens might cause IHD have not been extensively investigated, but likely involve coagulation and red blood cell attributes. Several haemostatic and thrombotic factors, such as thromboxane A2 [55, 56], endothelin-1 [57-59], nitric oxide [55, 60] and possibly thrombin [61, 62], may be driven by androgens and likely play a role in IHD [63-65]. Von Willebrand factor [66] and asymmetric dimethylarginine [67] are also modulated by statins and may cause IHD [4, 68], whether they are driven by testosterone is unknown. Several red blood cell attributes are affected by androgens, from reticulocytes to hematocrit [69, 70], but exactly which causes IHD is unclear, although reticulocytes are a possibility [71]. Currently, comprehensive genetic validation of these pathways is hampered by the lack of availability of large sex-specific GWAS of cytokines and coagulation factors.

Despite providing information that may be relevant to the performance of statins, and to the development of other therapies to protect against cardiovascular disease [72], some limitations of this study exist. First, valid instruments should fulfill three assumptions, i.e., relate strongly to the exposure, not be associated with potential confounders and satisfy the exclusion restriction assumption. The F-statistics were >10. Despite high conditional F-statistics for testosterone the conditional F-statistics for the effects of statin use were quite low and the Q-statistics for instrument validity were high suggesting pleiotropy, which we addressed by using multivariable MR-Egger. The associations with testosterone in women were not adjusted for factors, such as menopausal status, hormone use and history of oophorectomy, which could result in imprecision and weaker instruments. The SNPs used to predict effects of statin, PCSK9 inhibitors and ezetimibe use are well established [23], and in genes that harbor the target of each lipid modifier (*HMGCR, PCSK9* and *NCP1L1* respectively). Similarly, the SNPs predicting effects of the anti-inflammatory anakinra use have been validated as increasing IL-1Ra [22], and the SNP used to predict effects of tocilizumab use is well-established as affecting IL-6 and IL-6R [21]. Sex-specific genetic associations were used throughout with exception of the genetic prediction of effects of anakinra and tocilizumab use on IL1Ra and IL-6 respectively. However, inflammation operating on the reproductive axis would be expected to have sex-specific effects not sex-specific drivers. We selected between correlated SNPs based on p-values which is relatively arbitrary, and the estimates could be sensitive to the choice of SNPs. Repeating the analysis using a larger number of correlated SNPs, where possible, taking into account their correlation, gave a similar interpretation. MR studies can be confounded by population stratification. However, we used genetic associations from GWAS mainly comprising people of white British ancestry with genomic control. Functions of each SNP predicting the exposures are not all fully understood, so we cannot rule out the possibility that the SNPs are linked with IHD through other pathways although we used sensitivity analysis.

We used SNPs predicting testosterone, but not other exposures, obtained from the same study as the genetic effects on IHD. However given the estimates for testosterone were largely obtained from non-cases, the overlap unlikely introduced substantial bias [73]. Canalization, i.e., buffering of genetic factors during development, may occur however; whether it does so is unknown. Our findings, largely in Europeans, may not be applicable to other populations. However, causes are unlikely to act differently in different populations, although the causal mechanisms may not be as relevant in all settings [74]. The SNPs for effects of statin, PCSK9 inhibitors and ezetimibe use were selected for their relations with LDL-cholesterol and on functional grounds, assuming the lipid modifiers act by action on lipids, so it is possible that relevant SNPs might have been discarded if they work through other mechanisms independent of lipids. It is also possible that the SNPs for lipid modifiers might act via a different lipid trait, such as apoB [75]. The SNPs for effects of anakinra and tocilizumab use were similarly selected. Replication based on genetic instruments functionally relevant to all the exposures would be ideal. However, we used the most recent, published genetic instruments for testosterone [24]. Replication based on another large IHD GWAS, such as CARDIoGRAM [65], would be ideal, but it is less intensively genotyped than the UK Biobank and excludes the X chromosome, making it unsuitable. Lastly, Mendelian randomization assesses the lifelong effects of an endogenous exposure rather than short-term effects of an interventions assessed in an RCT, nevertheless our estimates for statins on IHD are comparable with meta-analyses of statin trials considering similar outcomes [13].

Taken together these complimentary findings for statins and anakinra raise the possibility that modulating testosterone, by whatever means, is a relevant feature for modulating IHD in men, with potential relevance to the development of new interventions, side-effects of existing interventions, re-purposing and appropriate use. Statins lowering testosterone could also be relevant to the muscle weakness or pain experienced on statins [76]. Recognition that statins lower testosterone might also provide greater impetus for investigation of their role in other relevant conditions, such as prostate cancer [77]. Conversely, statins and anakinra did not clearly affect testosterone in women (Table 1) nor did testosterone mediate the effect of statins on IHD in women (Table 3). These differences by sex highlight the need for sex-specific approaches to IHD prevention and management, specifically in terms of the use of statins and investigation more broadly of causes of IHD.

### Conclusion

Genetic variants corresponding to the effects of statins and anakinra use had opposite effects on testosterone and IHD in men, consistent with the effects of statins on IHD in men being partially mediated by testosterone. This insight that the pleiotropic effects of statins could be mediated by testosterone in men has implications for the use of existing interventions to prevent and treat IHD, the development of new interventions for IHD and the re-use of statins for other androgen related conditions. Genetic confirmation that anakinra raises testosterone suggests its use in rheumatoid arthritis might have cardiovascular side-effects, particularly in men. It also highlights the importance of considering whether vulnerability to major diseases and interventions to promote lifespan need to be sex-specific.

## Data Availability

This study only used data publicly available or available on request

## Contributorship

CMS initially planned, conducted, and reported the work described in this article. JVZ, SLAY and GML reviewed the first draft and suggested important additions to the design and content. JVZ additionally reviewed and checked the analysis plan and all the results. All authors reviewed and approved the final version. CMS is the guarantor. The corresponding author attests that all listed authors meet authorship criteria and that no others meeting the criteria have been omitted. CMS affirms that the manuscript is an honest, accurate, and transparent account of the study being reported; that no important aspects of the study have been omitted; and that any discrepancies from the study as planned have been explained.

